# Patient trajectories and risk factors for severe outcomes among persons hospitalized for COVID-19 in the Maryland/DC region

**DOI:** 10.1101/2020.05.24.20111864

**Authors:** Brian T. Garibaldi, Jacob Fiksel, John Muschelli, Matthew Robinson, Masoud Rouhizadeh, Paul Nagy, Josh H. Gray, Harsha Malapati, Mariam Ghobadi-Krueger, Timothy M. Niessen, Bo Soo Kim, Peter M. Hill, M. Shafeeq Ahmed, Eric D. Dobkin, Renee Blanding, Jennifer Abele, Bonnie Woods, Kenneth Harkness, David R. Thiemann, Mary G. Bowring, Aalok B. Shah, Mei-Cheng Wang, Karen Bandeen-Roche, Antony Rosen, Scott L. Zeger, Amita Gupta

## Abstract

**Background:** Risk factors for poor outcomes from COVID-19 are emerging among US cohorts, but patient trajectories during hospitalization ranging from mild-moderate, severe, and death and the factors associated with these outcomes have been underexplored.

**Methods:** We performed a cohort analysis of consecutive COVID-19 hospital admissions at 5 Johns Hopkins hospitals in the Baltimore/DC area between March 4 and April 24, 2020. Disease severity and outcomes were classified using the WHO COVID-19 disease severity ordinal scale. Cox proportional-hazards regressions were performed to assess relationships between demographics, clinical features and progression to severe disease or death.

**Results:** 832 COVID-19 patients were hospitalized; 633 (76.1%) were discharged, 113 (13.6%) died, and 85 (10.2%) remained hospitalized. Among those discharged, 518 (82%) had mild/moderate and 116 (18%) had severe illness. Mortality was statistically significantly associated with increasing age per 10 years (adjusted hazard ratio (aHR) 1.54; 95%CI 1.28-1.84), nursing home residence (aHR 2.13, 95%CI 1.41-3.23), Charlson comorbidity index (1.13; 95% CI 1.02-1.26), respiratory rate (aHR 1.13; 95%CI 1.09-1.17), D-dimer greater than 1mg/dL (aHR 2.79; 95% 1.53-5.09), and detectable troponin (aHR 2.79; 95%CI 1.53-5.09). In patients under 60, only male sex (aHR 1.7;95%CI 1.11-2.58), increasing body mass index (BMI) (aHR1.25 1.14-1.37), Charlson score (aHR 1.27; 1.1-1.46) and respiratory rate (aHR 1.16; 95%CI 1.13-1.2) were associated with severe illness or death.

**Conclusions:** A combination of demographic and clinical features on admission is strongly associated with progression to severe disease or death in a US cohort of COVID-19 patients. Younger patients have distinct risk factors for poor outcomes.

## INTRODUCTION

The first case of SARS-CoV-2 in the United States was identified January 20^th^, 2020 in a returned traveler from Wuhan, China.^1^ The US accounted for nearly a third of the world’s cases (1,577,758) and deaths (94,729) as of May 22, 2020.^2^ After infection with SARS-CoV-2, outcomes range from asymptomatic or mild illness to more severe illness and death.^3,4^ Age, sex, smoking, race, body mass index (BMI), and comorbidities such as hypertension and diabetes are important risk factors for severe outcomes, though to varying degrees. Elevated inflammatory markers and lymphopenia are also associated with severe outcomes in COVID-19 (the syndrome caused by SARS-CoV-2).^4-6^ While older age is one of the most important risk factors for hospitalization and death, it is increasingly recognized that younger persons may develop severe disease. Recently the CDC reported that 40% of hospitalizations in the US occurred among persons 20-50 years of age.^7^ Since most interventions under development to treat COVID-19 are likely to have a brief therapeutic window to prevent severe outcomes, it is critical to understand how factors on presentation to the hospital are related to disease progression. Such information is also important in determining appropriate levels of care as well as to guide discussions with patients and families. We performed a comprehensive analysis of the clinical features, patient trajectories and risk factors for progression to severe disease or death at the time of hospital admission.

## METHODS

### Study design and participants

This cohort study was conducted at five hospitals (Johns Hopkins Hospital, Baltimore, MD; Bayview Hospital, Baltimore, MD; Howard County General Hospital, Columbia, MD; Suburban Hospital, Bethesda, MD; Sibley Hospital, Washington DC) which comprise the Johns Hopkins Medicine System (JHM), a system licensed to operate 2,513 beds and 354 ICU beds serving approximately 7 million persons. The institutional review boards of these hospitals approved this study as minimal risk and waived requirement for informed consent. All patients consecutively admitted with confirmed SARS-CoV-2 infection by microbiological testing between March 4 and April 24, 2020 were included.

### Data collection

The primary data source was JH-CROWN: The COVID-19 PMAP Registry, which utilizes the Hopkins Precision Medicine Analytics Platform.^8^ Data in JH-CROWN include demographics, laboratory results, vital signs, respiratory events, medication administration, medical history, comorbid conditions, imaging, electrocardiogram results, and symptoms.

### Outcome Measures and Definitions

Our primary outcome was severe disease (including death), as defined by the WHO COVID-19 disease severity scale.^9^ This is an 8-point ordinal scale ranging from ambulatory (1=asymptomatic, 2=mild limitation in activity), to hospitalized with mild-moderate disease (3=room air, 4=nasal cannula or facemask oxygen), hospitalized with severe disease (5=high flow nasal canula (HFNC) or non-invasive positive pressure ventilation (NIPPV), 6=intubation and mechanical ventilation, 7=intubation and mechanical ventilation and other signs of organ failure (hemodialysis, vasopressors, extracorporeal membrane oxygenation (ECMO)), and 8=death. Peak COVID-19 severity score is reported as the maximum score during the observation period for individual patients. Multi-comorbidity burden was assessed using the Charlson Comorbidity Index (CCI).^10^

Diagnosis of COVID-19 was defined as detection of SARS-CoV-2 using any nucleic acid test with an Emergency Use Authorization from the US Food and Drug Administration. Samples predominantly included nasopharyngeal swabs and less commonly oropharyngeal swabs or bronchoalveolar lavage. Selection and frequency of other laboratory testing were determined by treating physicians. Natural Language Processing was used to identify presenting symptoms as described in the supplemental appendix (**Table S1**).

### Statistical Analyses

We estimated the cumulative incidence functions of death using the Aalen-Johansen estimator (CITE), with discharge and death as competing risks.^11^ To assess the association between patient characteristics and outcomes, a set of 24 demographic and clinical variables were selected based on clinical interest and knowledge. Missing values were imputed using multiple imputation by chained equations (MICE) with predictive mean matching,^12^ as implemented in the mice R package (version 3.7.0)^13,14^ with 10 rounds of multiple imputation. Cox proportional-hazard models were used to relate the risk of (i) dying and (ii) developing severe disease or dying to baseline patient characteristics.^15^ Four patients were discharged and then died. We censored their outcomes at time of discharge to minimize bias from lacking knowledge of deaths outside of the Hopkins system. We excluded the three pediatric patients (age <18 years) from the models. 92% of deaths occurred in patients over the age of 60, and almost half of those patients had a “do not resuscitate/no not intubate” (DNR/DNI) order within 24 hours of admission. We used a cox proportional-hazards model to relate the risk of severe illness or death to baseline characteristics in patients under the age of 60 in order to capture the risks of severe illness in that population.

Models were initially built adding variables in categorized “blocks” (e.g. “demographic”) to protect against overfitting. For the composite outcome of severe disease or death, findings were equivalent to those from a model including all covariates. For other models, further variable selection was warranted when including multiple covariate blocks simultaneously. Here, we fit cause-specific proportional-hazards models regularized with an elastic net penalty, as implemented in the glmnet R package (version 3.0.2).^16^ The elastic net model was run on each of the 10 imputed datasets, and variables with non-zero coefficients in at least half of the models were chosen for the final model,^17^ which was again run on each of the 10 imputed datasets. Demographic variables were forced to be in the models. No variable selection was done for the time to composite severe outcome or death model, as there were a sufficient number of events to allow for a larger model. Standard error estimates were computed using Rubin’s rules (Rubin, 2004),^18^ as implemented in the mice R package. All analysis was done using R Version 3.6.2.^19^

## RESULTS

### Study population

A total of 832 adult and pediatric patients were admitted with confirmed SARS-CoV-2 infection from March 4 to April 24 (**Figure 1a**). The median age was 63 years (IQR 49,75, range 1-108), 443 (53%) were male, median BMI was 29 kg/m^2^ (IQR 25,34, range 15.2-74.9), 333 (40%) were African American, 134 (16%) were Latinx and 146 (18%) were nursing home residents. **Table 1** summarizes demographics, comorbidities, presenting symptoms, laboratory data and vital signs at presentation to the hospital for all patients, and divided into disease categories of mild-moderate, severe and death.

**Figure 1.**
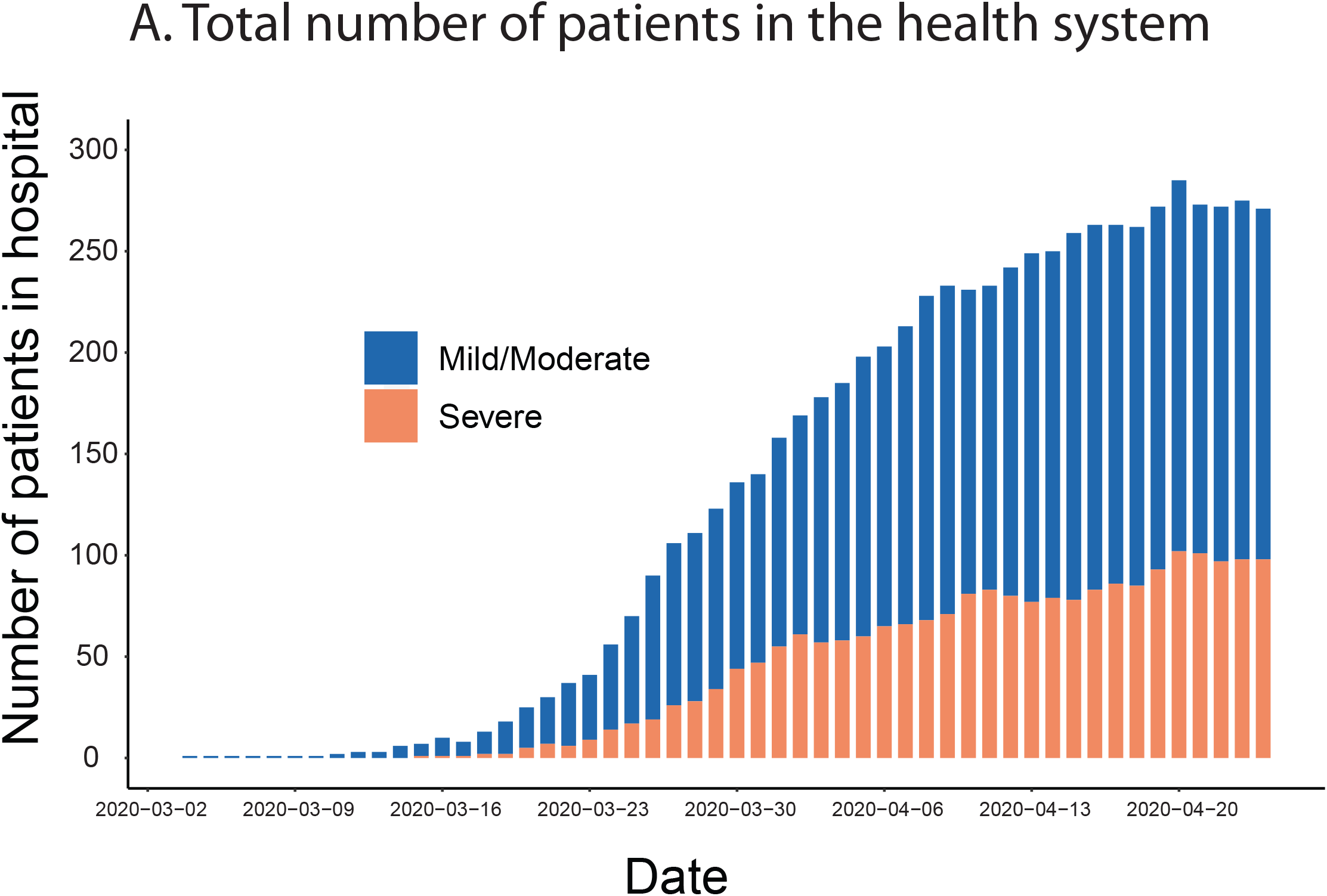
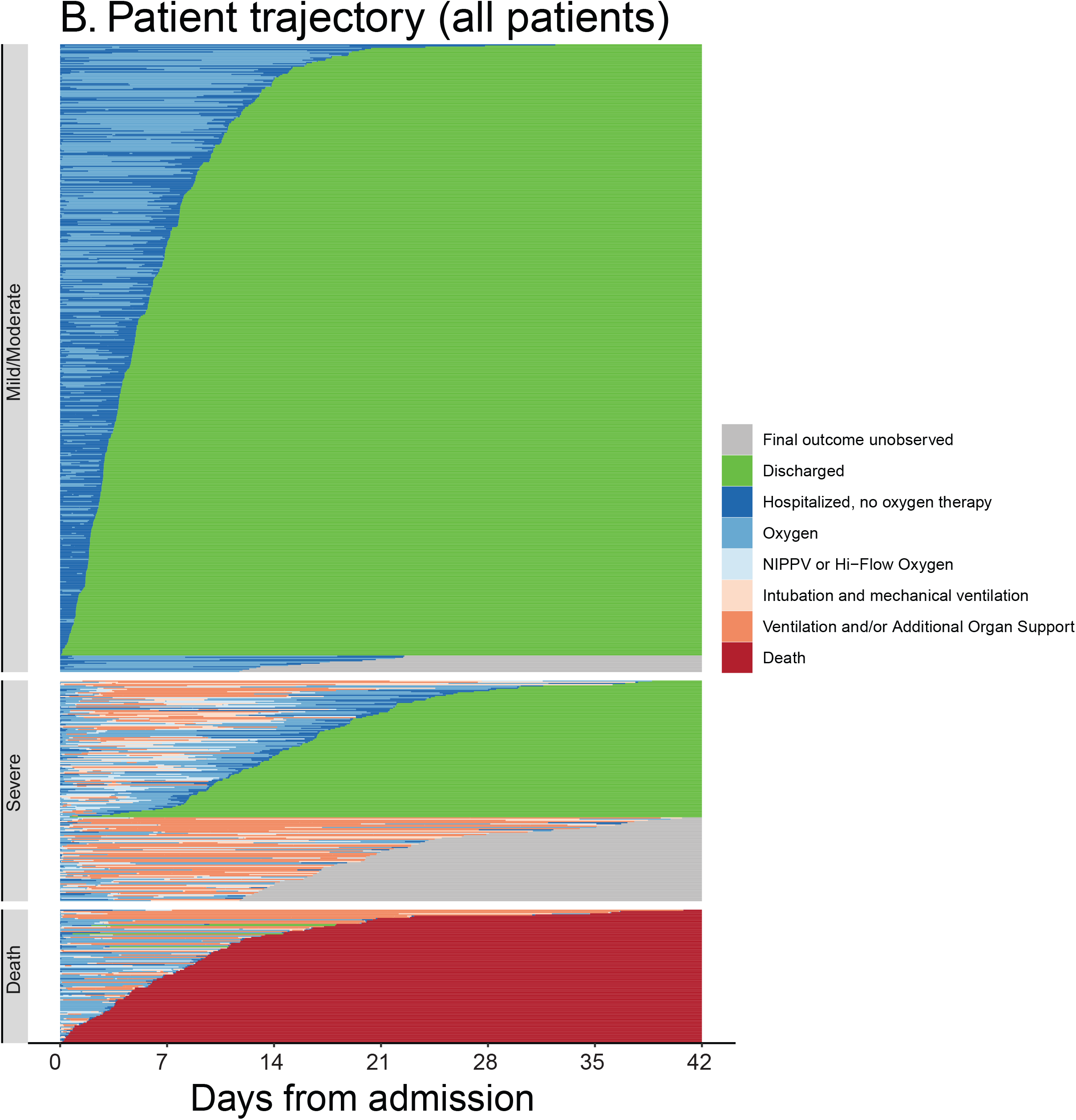
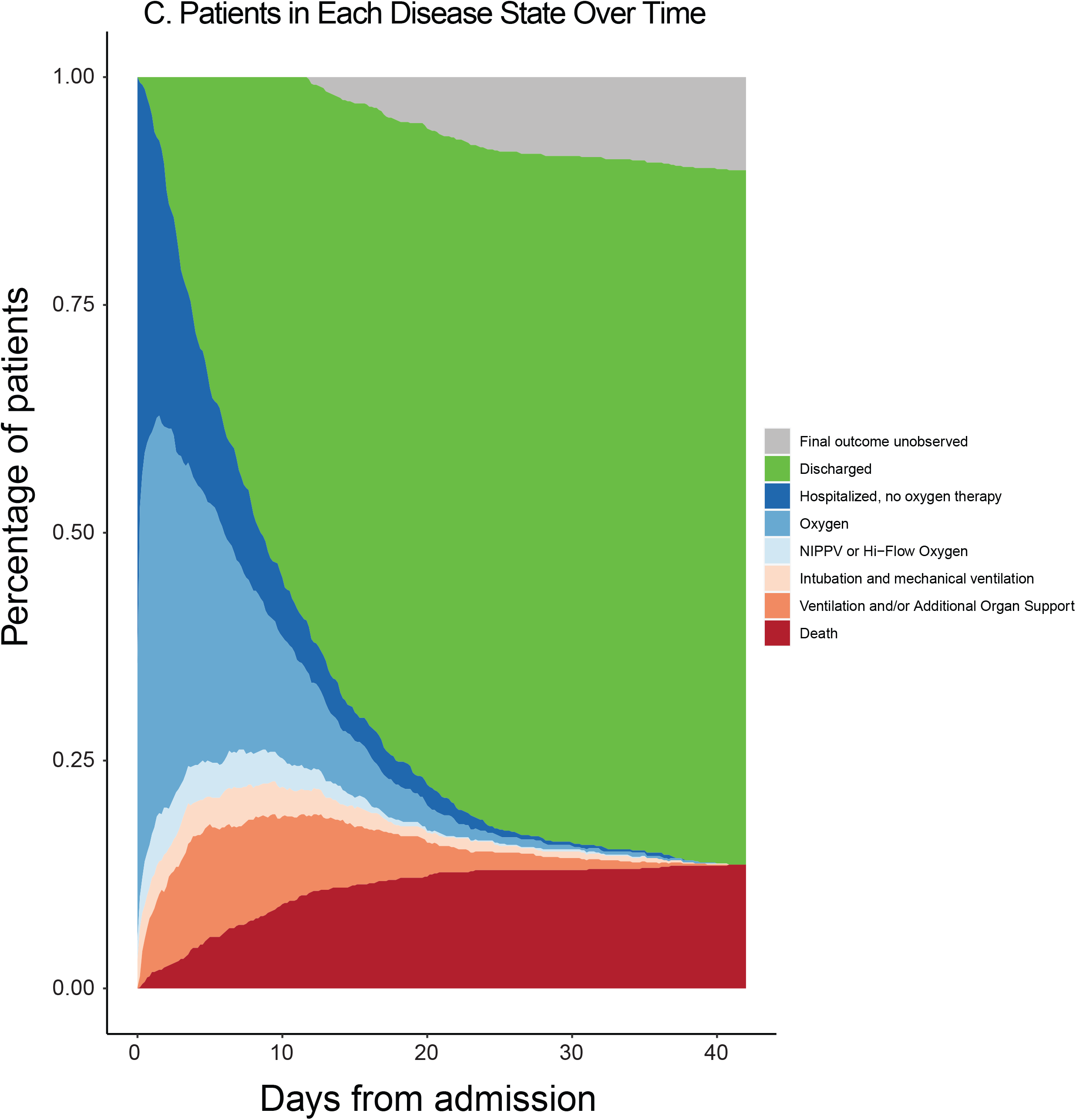
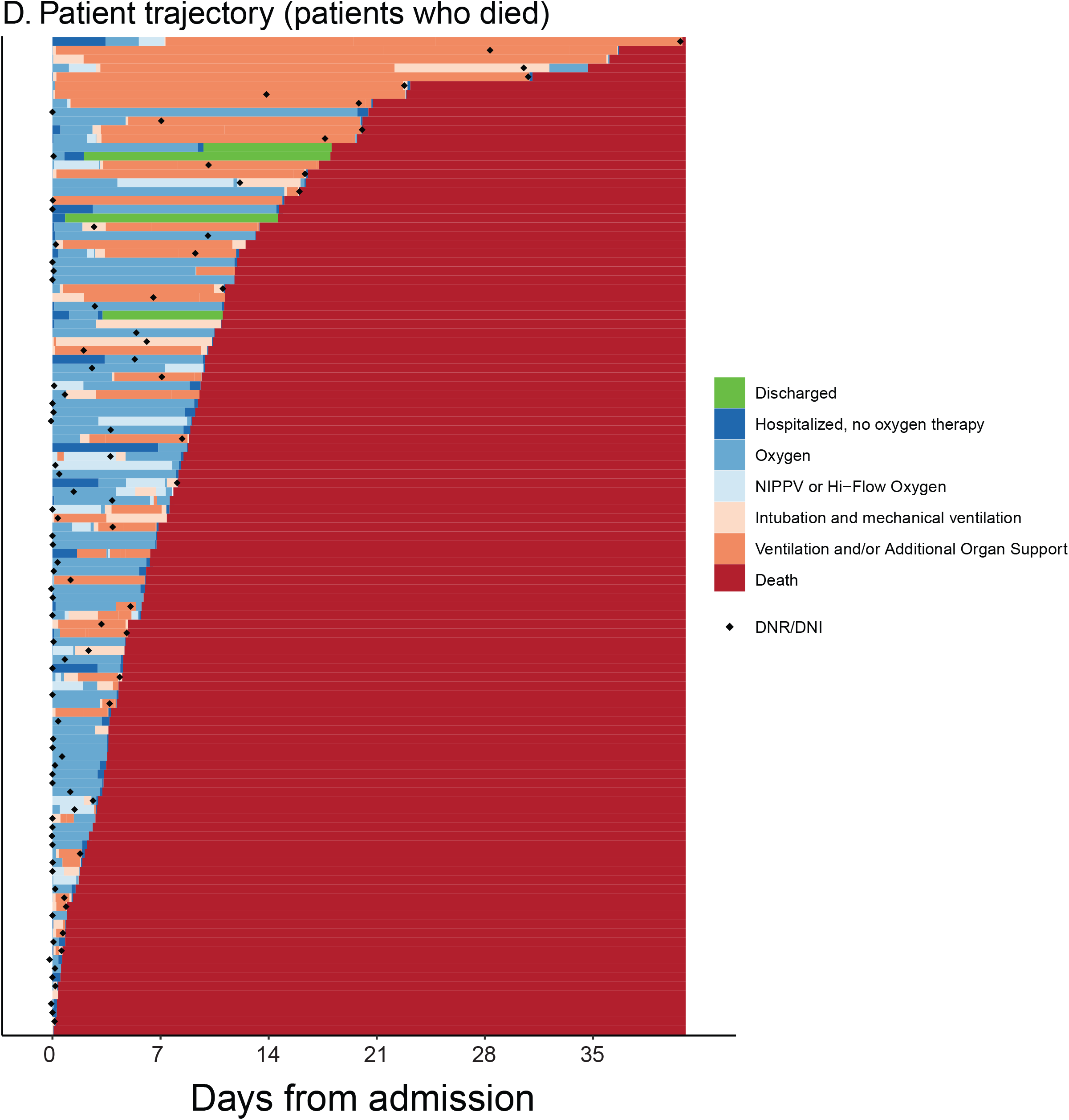
Disease course and trajectories for patients admitted with COVID-19. **1a. COVID-19 Admissions and census for the Johns Hopkins Medical System**. The Johns Hopkins Medical System in the Baltimore-Washington DC region is comprised of 5 hospitals with 2,513 beds, including 354 ICU beds and serves a population of 6.7 million. The first case was reported on March 3, 2020. The last patients in this cohort were admitted on April 24; data was censored on May 5, 2020. The figure shows the number of COVID-19 positive patients admitted across the health system each day during the study period, and the numbers with mild disease compared to severe disease. **1b. Patient trajectory according to WHO ordinal scale**. Patient trajectories illustrating WHO COVID-19 disease stage transitions plotted by days from admission and grouped by peak WHO disease stage with individual colors showing specific transitions. Patients who have been discharged (represented in bright green), patients who remain hospitalized and have not yet achieved a final outcome (represented in gray), and patients who have died (represented in red) are illustrated. Each horizontal line is an individual patient. The median length of stay for patients with mild/moderate disease was 4.8 days (IQR 2.56,8.08). The median length of stay for patients with severe disease was 15 days (IQR 10.2,20.58). The median length of stay for those who died was 6.8 days (IQR 3.3, 11.2). Among 199 patients who were mechanically ventilated, 74 (37.2%) were discharged, 63 (31.7%) died and 62 (31.2%) remain hospitalized, of which 43 (21.6%) remained on the ventilator. **1c. Proportion of hospitalized patients in each WHO COVID-19 disease stage and discharged by days from admission**. This figure illustrates the fraction of the 832 patients who are in each WHO disease state at a given time post admission to the hospital. Note that both discharge (green) and death (red) are cumulative. The percentage of patients for whom their WHO disease stage is unknown after a given time due to administrative censoring is represented in grey. **1d. Patient trajectories among those who died**. This figure illustrates the trajectory of each patient who died. Each horizontal line represents a single patient. The black diamonds represent the day that a code status including either a do not resuscitate (DNR) or do not intubate (DNI) was entered into the medical record. Fifty-five patients (49%) had a DNR/DNI order placed within 24 hours of admission (n=113 deaths).

**Table 1.** Baseline Demographic and Clinical Characteristics by Observed Outcome.

| Characteristic | Overall, N = 832 | Final outcome observed, N=747 |  |  | Final outcome not yet observed, N=85 |  |
| --- | --- | --- | --- | --- | --- | --- |
|  |  | Mild/Moderate (N = 518 <sup>a</sup>) | Severe (N = 116 <sup>b</sup>) | Death (N = 113) | Mild/Moderate (N = 14) | Severe (N = 71) |
| Demographics<sup>c</sup> |  |  |  |  |  |  |
| Median age (IQR) | 63 (49, 75) | 60 (45, 72) | 56 (49, 70) | 78 (68, 86) | 74 (61, 85) | 64 (54, 75) |
| Female | 389 (46.8%) | 256 (49.4%) | 50 (43.1%) | 47 (41.6%) | 6 (42.9%) | 30 (42.3%) |
| Pregnant | 13 (1.6%) | 11 (2.1%) | 2 (1.7%) | 0 (0.0%) | 0 (0.0%) | 0 (0.0%) |
| Race and ethnicity |  |  |  |  |  |  |
| Asian | 48 (5.8%) | 32 (6.2%) | 3 (2.6%) | 9 (8.0%) | 0 (0.0%) | 4 (5.6%) |
| Black | 333 (40.4%) | 206 (40.2%) | 43 (37.4%) | 43 (38.4%) | 6 (42.9%) | 35 (49.3%) |
| Hispanic | 134 (16.3%) | 90 (17.6%) | 23 (20.0%) | 6 (5.4%) | 1 (7.1%) | 14 (19.7%) |
| Caucasian, Non-Hispanic | 266 (32.3%) | 155 (30.3%) | 37 (32.2%) | 52 (46.4%) | 7 (50.0%) | 15 (21.1%) |
| Other/multiracial | 43 (5.2%) | 29 (5.7%) | 9 (7.8%) | 2 (1.8%) | 0 (0.0%) | 3 (4.2%) |
| Admitted from skilled nursing facility | 146 (17.7%) | 65 (12.6%) | 13 (11.2%) | 55 (49.1%) | 0 (0.0%) | 13 (18.6%) |
| Alcohol use | 188 (25.9%) | 128 (27.7%) | 31 (29.5%) | 15 (16.3%) | 2 (15.4%) | 12 (22.2%) |
| Smoking |  |  |  |  |  |  |
| Current Smoker | 46 (6.3%) | 24 (5.1%) | 12 (11.1%) | 6 (7.3%) | 1 (8.3%) | 3 (5.3%) |
| Former Smoker | 188 (25.8%) | 102 (21.7%) | 27 (25.0%) | 36 (43.9%) | 5 (41.7%) | 18 (31.6%) |
| Median BMI (kg/m<sup>2</sup>; IQR) | 29 (25, 34) | 28 (25, 34) | 31 (27, 35) | 27 (24, 33) | 26 (23, 31) | 33 (27, 37) |
| DNR/DNI in first 24 hrs of admission | 124 (14.9%) | 56 (10.8%) | 8 (6.9%) | 55 (48.7%) | 2 (14.3%) | 3 (4.2%) |
| Comorbidities |  |  |  |  |  |  |
| Hypertension | 410 (49.3%) | 234 (45.2%) | 50 (43.1%) | 78 (69.0%) | 12 (85.7%) | 36 (50.7%) |
| Coronary Artery Disease | 270 (32.5%) | 126 (24.3%) | 41 (35.3%) | 61 (54.0%) | 10 (71.4%) | 32 (45.1%) |
| Congestive heart failure | 127 (15.3%) | 42 (8.1%) | 25 (21.6%) | 33 (29.2%) | 7 (50.0%) | 20 (28.2%) |
| Chronic Kidney Disease | 107 (12.9%) | 46 (8.9%) | 13 (11.2%) | 34 (30.1%) | 4 (28.6%) | 10 (14.1%) |
| Diabetes | 261 (31.4%) | 146 (28.2%) | 34 (29.3%) | 38 (33.6%) | 6 (42.9%) | 37 (52.1%) |
| Asthma | 87 (10.5%) | 58 (11.2%) | 16 (13.8%) | 8 (7.1%) | 0 (0.0%) | 5 (7.0%) |
| COPD/Chronic Lung Disease | 168 (20.2%) | 101 (19.5%) | 26 (22.4%) | 27 (23.9%) | 6 (42.9%) | 8 (11.3%) |
| Cancer | 90 (10.8%) | 46 (8.9%) | 12 (10.3%) | 20 (17.7%) | 2 (14.3%) | 10 (14.1%) |
| Liver disease | 36 (4.3%) | 18 (3.5%) | 11 (9.5%) | 3 (2.7%) | 1 (7.1%) | 3 (4.2%) |
| Immunosuppressed | 22 (2.6%) | 15 (2.9%) | 4 (3.4%) | 2 (1.8%) | 0 (0.0%) | 1 (1.4%) |
| AIDS/HIV | 11 (1.3%) | 6 (1.2%) | 3 (2.6%) | 1 (0.9%) | 0 (0.0%) | 1 (1.4%) |
| Transplant | 20 (2.4%) | 15 (2.9%) | 3 (2.6%) | 2 (1.8%) | 0 (0.0%) | 0 (0.0%) |
| Charlson comorbidity index |  |  |  |  |  |  |
| 0 | 288 (34.6%) | 210 (40.5%) | 42 (36.2%) | 17 (15.0%) | 1 (7.1%) | 18 (25.4%) |
| 1-2 | 379 (45.6%) | 235 (45.4%) | 51 (44.0%) | 53 (46.9%) | 6 (42.9%) | 34 (47.9%) |
| 3-4 | 121 (14.5%) | 65 (12.5%) | 13 (11.2%) | 27 (23.9%) | 3 (21.4%) | 13 (18.3%) |
| >=5 | 44 (5.3%) | 8 (1.5%) | 10 (8.6%) | 16 (14.2%) | 4 (28.6%) | 6 (8.5%) |
| **Vital Signs within 24 hrs of admission, Median (IQR)<sup>d</sup>** |  |  |  |  |  |  |
| Respiratory rate (breaths/min) | 20.0 (18.0, 22.0) | 18.5 (18.0, 20.0) | 22.0 (19.8, 25.5) | 24.0 (20.0, 27.6) | 18.5 (18.0, 19.0) | 22.5 (19.0, 27.5) |
| Pulse (beats/min) | 84 (75, 95) | 83 (74, 93) | 87 (78, 96) | 86 (73, 98) | 86 (76, 90) | 85 (76, 96) |
| Temperature Maximum (degrees Celsius) | 37.80 (37.20, 38.60) | 37.70 (37.20, 38.40) | 38.30 (37.60, 39.20) | 37.90 (37.20, 38.70) | 37.75 (37.10, 38.08) | 37.90 (37.30, 38.75) |
| FiO2 | 0.28 (0.20, 0.36) | 0.20 (0.20, 0.28) | 0.36 (0.28, 0.50) | 0.44 (0.30, 0.80) | 0.20 (0.20, 0.27) | 0.44 (0.28, 0.70) |
| SaO2/FiO2 | 371 (274, 480) | 472 (348, 482) | 251 (206, 343) | 220 (121, 325) | 475 (366, 485) | 227 (141, 328) |
| Mean arterial blood pressure (mmHg) | 86 (80, 94) | 87 (81, 95) | 84 (77, 94) | 82 (76, 88) | 86 (82, 89) | 84 (77, 91) |
| Intubated in the first 24 hrs | 70 (8.5%) | 0 (0.0%) | 22 (19.0%) | 24 (21.4%) | 0 (0.0%) | 24 (33.8%) |
| NIPPV or HFNC in first 24 hrs | 76 (9.2%) | 2 (0.4%) | 33 (28.4%) | 18 (16.1%) | 0 (0.0%) | 23 (32.4%) |
| **Presenting Symptoms** |  |  |  |  |  |  |
| Cough | 649 (78.0%) | 413 (79.7%) | 105 (90.5%) | 71 (62.8%) | 11 (78.6%) | 49 (69.0%) |
| Shortness of breath | 689 (82.8%) | 403 (77.8%) | 107 (92.2%) | 102 (90.3%) | 11 (78.6%) | 66 (93.0%) |
| Fever | 691 (83.1%) | 429 (82.8%) | 108 (93.1%) | 88 (77.9%) | 12 (85.7%) | 54 (76.1%) |
| Chills | 343 (41.2%) | 240 (46.3%) | 59 (50.9%) | 19 (16.8%) | 4 (28.6%) | 21 (29.6%) |
| Muscle pain | 353 (42.4%) | 252 (48.6%) | 55 (47.4%) | 16 (14.2%) | 3 (21.4%) | 27 (38.0%) |
| Headache | 156 (18.8%) | 123 (23.7%) | 23 (19.8%) | 2 (1.8%) | 2 (14.3%) | 6 (8.5%) |
| Sore throat | 191 (23.0%) | 138 (26.6%) | 27 (23.3%) | 12 (10.6%) | 3 (21.4%) | 11 (15.5%) |
| New loss of taste or smell | 134 (16.1%) | 103 (19.9%) | 18 (15.5%) | 4 (3.5%) | 2 (14.3%) | 7 (9.9%) |
| Abdominal pain | 148 (17.8%) | 103 (19.9%) | 22 (19.0%) | 14 (12.4%) | 2 (14.3%) | 7 (9.9%) |
| Diarrhea | 296 (35.6%) | 192 (37.1%) | 54 (46.6%) | 24 (21.2%) | 3 (21.4%) | 23 (32.4%) |
| Vomiting | 305 (36.7%) | 204 (39.4%) | 50 (43.1%) | 23 (20.4%) | 5 (35.7%) | 23 (32.4%) |
| Appetite loss | 254 (30.5%) | 179 (34.6%) | 32 (27.6%) | 24 (21.2%) | 6 (42.9%) | 13 (18.3%) |
| Fatigue | 572 (68.8%) | 372 (71.8%) | 79 (68.1%) | 61 (54.0%) | 12 (85.7%) | 48 (67.6%) |
| **Laboratory Data in first 24hrs (Median, IQR)<sup>e</sup>** |  |  |  |  |  |  |
| White blood cell count (K/cu mm) | 6.7 (4.9, 9.1) | 6.4 (4.7, 8.3) | 6.7 (5.1, 9.7) | 8.1 (5.8, 10.9) | 7.0 (4.8, 9.1) | 7.0 (6.1, 9.3) |
| White blood cell count range (K/cu mm) |  |  |  |  |  |  |
| <4 | 101 (12.2%) | 74 (14.3%) | 10 (8.6%) | 10 (8.8%) | 2 (14.3%) | 5 (7.0%) |
| >12 | 93 (11.2%) | 43 (8.3%) | 17 (14.7%) | 22 (19.5%) | 1 (7.1%) | 10 (14.1%) |
| 4-12 | 637 (76.7%) | 400 (77.4%) | 89 (76.7%) | 81 (71.7%) | 11 (78.6%) | 56 (78.9%) |
| Absolute Lymphocyte Count (K/cu mm) | 0.99 (0.69, 1.43) | 1.07 (0.76, 1.54) | 0.93 (0.66, 1.29) | 0.79 (0.52, 1.13) | 1.05 (0.64, 1.33) | 0.84 (0.57, 1.20) |
| Absolute lymphocyte count range (K/cu mm) |  |  |  |  |  |  |
| <0.8 | 281 (34.2%) | 142 (27.9%) | 46 (39.7%) | 56 (50.0%) | 5 (35.7%) | 32 (45.7%) |
| >=0.8 | 540 (65.8%) | 367 (72.1%) | 70 (60.3%) | 56 (50.0%) | 9 (64.3%) | 38 (54.3%) |
| Hemoglobin (g/dL) | 13.10 (11.60, 14.30) | 13.10 (11.90, 14.40) | 13.40 (11.90, 14.70) | 12.80 (10.70, 14.30) | 11.85 (10.68, 13.05) | 12.30 (10.75, 13.65) |
| Platelet Count (K/cu mm) | 204 (157, 261) | 203 (158, 259) | 206 (164, 258) | 188 (141, 246) | 247 (170, 294) | 226 (160, 286) |
| Albumin (g/dL) | 3.70 (3.30, 4.10) | 3.80 (3.50, 4.20) | 3.60 (3.30, 3.90) | 3.50 (3.00, 3.80) | 3.15 (2.60, 3.98) | 3.40 (3.00, 3.70) |
| ALT (U/L) | 29 (19, 48) | 29 (18, 47) | 35 (24, 51) | 30 (18, 42) | 20 (14, 44) | 28 (18, 53) |
| AST (U/L) | 39 (27, 58) | 36 (24, 53) | 46 (33, 64) | 42 (30, 72) | 29 (21, 48) | 43 (34, 77) |
| Bilirubin (mg/dL) | 0.50 (0.30, 0.60) | 0.40 (0.30, 0.60) | 0.50 (0.40, 0.65) | 0.50 (0.40, 0.80) | 0.40 (0.20, 0.67) | 0.50 (0.30, 0.60) |
| Creatinine (mg/dL) | 1.00 (0.80, 1.50) | 0.97 (0.80, 1.30) | 1.01 (0.80, 1.42) | 1.60 (1.10, 2.20) | 1.00 (0.80, 2.15) | 1.40 (0.90, 2.50) |
| GFR (mL/min/1.73sqm) | 74 (44, 97) | 83 (55, 102) | 72 (52, 96) | 38 (24, 66) | 64 (28, 90) | 46 (24, 88) |
| CRP (mg/dL) | 8 (3, 14) | 6 (2, 11) | 11 (6, 17) | 13 (7, 19) | 4 (2, 9) | 12 (8, 21) |
| Procalcitonin (ng/ml) | 0.20 (0.10, 0.55) | 0.12 (0.10, 0.31) | 0.21 (0.13, 0.49) | 0.56 (0.21, 2.11) | 0.20 (0.14, 0.44) | 0.44 (0.20, 1.17) |
| LDH (U/L) | 340 (250, 484) | 300 (221, 413) | 402 (317, 508) | 469 (318, 628) | 237 (168, 310) | 474 (348, 616) |
| D-Dimer (mg/L FEU) | 0.93 (0.52, 1.83) | 0.75 (0.45, 1.40) | 0.88 (0.50, 1.32) | 2.06 (1.14, 4.00) | 1.82 (0.85, 2.88) | 1.29 (0.75, 2.32) |
| D-Dimer range (mg/L FEU) |  |  |  |  |  |  |
| 0-0.5 | 141 (24.2%) | 105 (30.4%) | 25 (26.9%) | 5 (6.7%) | 0 (0.0%) | 6 (9.8%) |
| 0.5-1 | 166 (28.5%) | 107 (31.0%) | 29 (31.2%) | 10 (13.3%) | 3 (33.3%) | 17 (27.9%) |
| >1 | 276 (47.3%) | 133 (38.6%) | 39 (41.9%) | 60 (80.0%) | 6 (66.7%) | 38 (62.3%) |
| Fibrinogen (mg/dL) | 511 (397, 611) | 482 (375, 586) | 616 (576, 735) | 484 (352, 632) | 445 (283, 445) | 548 (460, 641) |
| Ferritin (ng/ml) | 590 (280, 1103) | 506 (216, 883) | 681 (336, 1301) | 871 (454, 1881) | 328 (127, 570) | 830 (448, 1372) |
| IL6 (pg/ml) | 49 (19, 122) | 23 (13, 53) | 80 (38, 134) | 154 (103, 171) | 54 (31, 70) | 151 (77, 354) |
| Hemoglobin A1c (%) | 6.80 (5.90, 8.63) | 6.70 (5.80, 8.85) | 6.80 (6.00, 7.90) | 7.35 (6.70, 8.23) | 5.60 (5.55, 6.35) | 7.50 (6.00, 9.10) |
| Troponin above limit of detection | 202 (29.6%) | 70 (17.2%) | 26 (26.3%) | 70 (70.0%) | 4 (40.0%) | 32 (49.2%) |
| Pro-BNP (pg/ml) | 208 (45, 946) | 113 (24, 468) | 206 (72, 758) | 1014 (218, 4156) | 1265 (565, 1766) | 487 (108, 1356) |

<sup>a</sup>Mild to moderate include patients with WHO ordinal score of 3 (not on oxygen) and 4 (on nasal cannula or face-mask oxygen). <sup>b</sup>Severe includes patients with WHO ordinal score of 5 (high-flow nasal cannula or non-invasive positive pressure ventilation), 6 (intubation and mechanical ventilation), and 7 (intubated, mechanical ventilation and other signs of organ failure including ECMO, hemodialysis or vasopressors). <sup>c</sup>Data was missing for race and ethnicity data in 8 patients, alcohol use in 106 patients, smoking history in 103 patients, and BMI for 16 patients. <sup>d</sup>Data was missing for admission vital signs for 11 patients, temperature for 13 patients, positive pressure ventilation use for 10 patients, and SaO<sub>2</sub>/FiO<sub>2</sub> for 15 patients. <sup>e</sup>Data was missing for WBC for 1 patient, ALC for 11 patients, hemoglobin for 1 patient, platelet count for 2 patients, albumin for 13 patients, ALT for 15 patients, AST for 40 patients, bilirubin for 13 patients, creatinine for 2 patients, GFR for 4 patients, CRP for 220 patients, procalcitonin for 382 patients, LDH for 372 patients, D-Dimer for 249 patients, fibrinogen for 603 patients, ferritin for 271 patients, IL-6 for 651 patients, hemoglobin A1c for 580 patients, troponin for 150 patients, and Pro-BNP for 401 patients.

### Clinical Course and key characteristics by disease outcome

Among the 832 hospitalized patients, 633 (76.1%) were discharged, 113 (13.6%) died, and 85 (10.2%) remained hospitalized at day of censoring (May 4, 2020). Patient trajectories and WHO illness state during hospitalization are shown in **Figures 1b-d**. Among those discharged, the peak illness state was as follows: 518 (81.8%) were mild/moderate; 116 (18.3%) were severe with 88 (11.8%) requiring HFNC oxygen or NIPPV and 74 (9.9%) requiring invasive mechanical ventilation. Among 199 patients who were mechanically ventilated, 74 (37.2%) were discharged, 63 (31.7%) died and 62 (31.2%) remain hospitalized, of which 43 (21.6%) remained on the ventilator. Six patients were placed on ECMO; two died, three were successfully decannulated and one remained on ECMO. 637 (76.6%) patients received systemic antibiotics, 384 (46.2%) received hydroxychloroquine, 121 (14.5%) received systemic corticosteroids, 151 (18.1%) received ACE-inhibitors or ARBs, 39 (4.7%) received tocilizumab, and 14 (1.7%) were enrolled in clinical trials (**Table S2**).

Among the 113 patients who died, 55 (48.7%) developed shock requiring vasopressors, 46 (40.7%) developed acute renal failure, 17 (15.0%) required new hemodialysis, 12 (10.6%) developed bacteremia, 10 (8.8%) developed a ventilator-associated pneumonia, and 4 (3.5%) developed fungemia. Three (2.6%) patients had an ischemic stroke, 4 (3.5%) had an intracranial hemorrhage, 2 (1.8%) had a pulmonary embolism, and 2 (1.8%) had a deep venous thrombosis. Most (n=89, 78.8%) had a DNR/DNI order placed during their admission, with 55 (48.7%) having a DNR/DNI order within 24 hours of admission.

The overall median length of stay was 6.1 days (IQR 2.6, 10.8). Those who died, and those who had severe illness had a longer median length of stay compared to those with mild/moderate illness (mild-moderate - 4.8 days [IQR 2.6,8]; severe - 15 days [IQR 10.2,20.6]; death - 6.8 days [IQR 3.3,11.2]).

### Factors on admission associated with severe illness or death

As shown in **Table 1, Figure 2 and Figure 3**, several characteristics distinguished peak illness states. Increasing age (aHR 1.54 per 10-year increase; 95% CI 1.28-1.84), admission from a nursing home (aHR 2.13; 95% CI 1.41-3.23) and increasing CCI (aHR 1.13; 95%CI 1.02-1.26) were independently associated with death (**Table 2**). Respiratory rate (aHR 1.13 per increase of 1 over 18; 95% CI 1.09-1.17), a detectable troponin (aHR 2.49; 95%CI 1.5-4.14) and D-dimer greater than 1 mg/dL (aHR 2.79; 95%CI 1.53-5.09) were also significantly associated with death. An additional model including SaO2/FiO2 is shown in **Table S3**. SaO2/FiO2 ratio was significantly associated with death (aHR 1.35 per decrease of 50 below 375; 95%CI 1.16-1.56); all variables in the initial model retained their significance.

**Figure 2.**
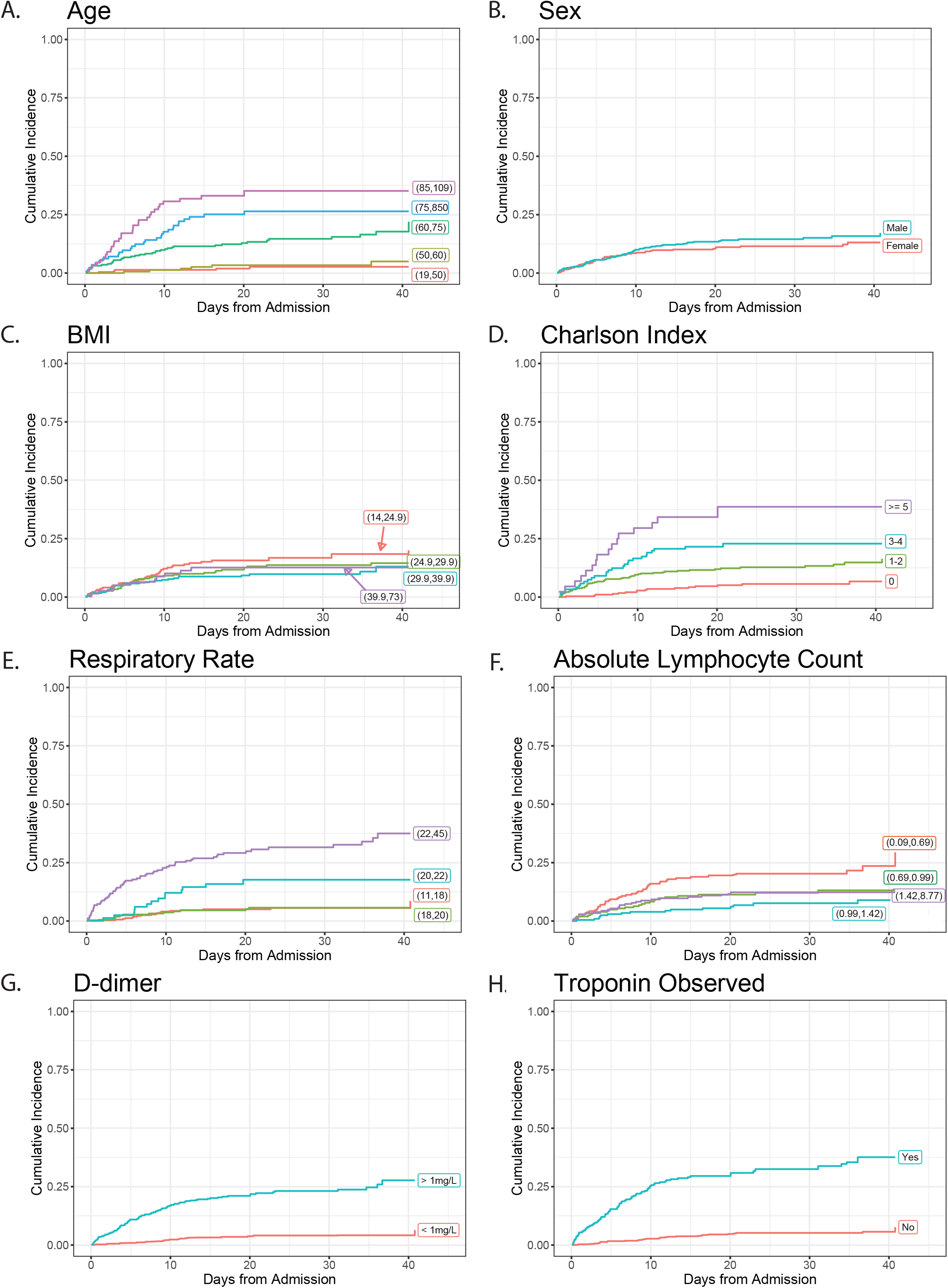
Cumulative incidence curves for death for key patient characteristics. This figure shows the cumulative incidence curves for death for key patient characteristics. In multivariate analyses increasing age (A), increasing BMI (C), Charlson Comorbidity Index (D), increasing respiratory rate greater (E), a D-dimer greater than 1 mg/L (G) and a detectable troponin (H) and were significantly associated with death. Decreasing absolute lymphocyte count (F) was significantly associated with progression to severe disease or death in a separate multivariate analysis. Male sex (B) was associated with severe disease or death in patients under the age of 60.

**Figure 3.**
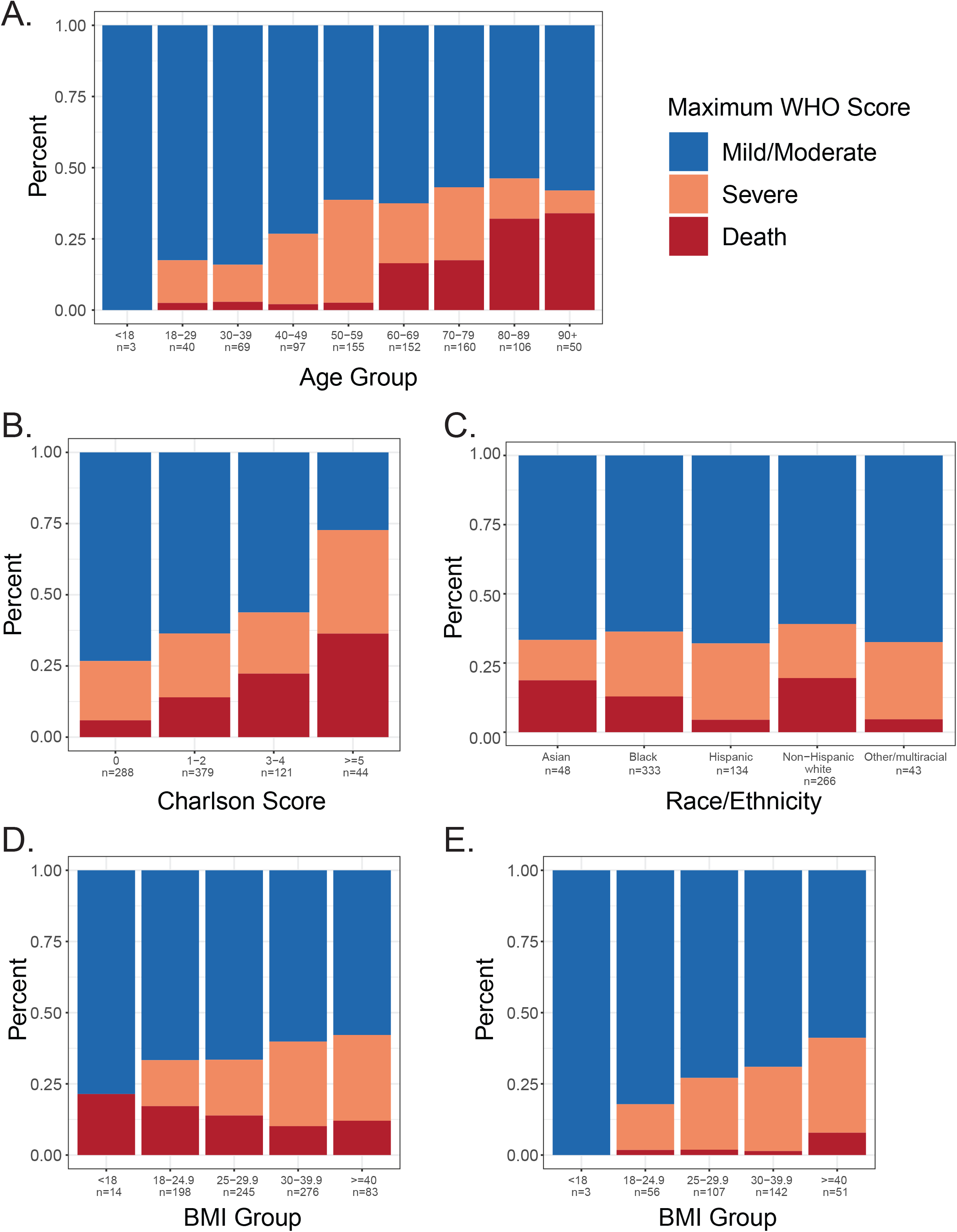
Disease Severity Grouped by Specific Characteristics. This figure shows the distribution of WHO disease severity states grouped according to specific characteristics. A. WHO maximum disease state grouped by age (n=832). B. WHO maximum disease state grouped by Charlson score (n=832). C. WHO maximum disease state grouped by race/ethnicity (n=832). D. WHO maximum disease state grouped by BMI (n=832). E. WHO maximum disease state grouped by BMI for patients under 60 (n=364).

**Table 2:** Association of baseline clinical characteristics and severe and death outcomes.

| Clinical characteristic† | Severe outcome or death | Severe outcome or death, age < 60 | Death |
| --- | --- | --- | --- |
|  | Hazard ratio (95% CI)‡<br>(N = 829,<br>296/4012 events/person-days<br>observed) | Hazard ratio (95% CI)‡<br>(N=361,<br>104/1417 events/person-days<br>observed) | Hazard ratio (95% CI)<br>(N=829,<br>109/7635 events/person-days<br>observed) |
| Age (increments of 10 years) | 0.94 (0.86-1.03) | 1.21 (0.96-1.51) | 1.54 (1.28-1.84)* |
| Admission from nursing home | 1.01 (0.74-1.37) |  | 2.13 (1.41-3.23)* |
| Male sex | 1.05 (0.8-1.37) | 1.7 (1.11-2.58)* | 1.24 (0.82-1.87) |
| Body mass index (increments of 5 kg/m2) | 1.19 (1.11-1.28)* | 1.25 (1.14-1.37)* | 1.11 (0.98-1.25) |
| Non-white race/ethnicity | 1.04 (0.79-1.37) | 1.07 (0.6-1.89) | 0.94 (0.61-1.45) |
| Charlson comorbidity score | 1.02 (0.94-1.12) | 1.27 (1.1-1.46)* | 1.13 (1.02-1.26)* |
| Respiratory symptoms | 2.11 (1.13-3.94)* |  |  |
| Gastrointestinal symptoms | 1.22 (0.94-1.57) |  |  |
| Constitutional symptoms | 0.56 (0.36-0.87)* |  |  |
| Loss of taste and/or smell | 0.76 (0.5-1.16) |  |  |
| Respiratory rate < 18 breaths per minute | 0.84 (0.7-1.01) | 0.79 (0.62-1)* | 0.83 (0.63-1.09) |
| Respiratory rate > 18 breaths per minute | 1.12 (1.1-1.15)* | 1.16 (1.13-1.2)* | 1.13 (1.09-1.17)* |
| Pulse (increments of 10) | 1 (0.92-1.08) |  |  |
| Fever (Temperature >38.4° Celsius) | 1.77 (1.36-2.32)* |  |  |
| White blood cell count (log) | 1 (0.77-1.3) |  |  |
| Absolute lymphocyte count (log scale) | 0.73 (0.58-0.91)* |  |  |
| Hemoglobin | 1.05 (0.98-1.12) |  |  |
| Albumin (every decrease of 0.5 g/dL up to 4g/dL) | 1.30 (1.15-1.47)* |  |  |
| Alanine aminotransferase (log scale) | 0.96 (0.81-1.14) |  |  |
| Glomerular filtration rate (increments of 10 ml/min) | 0.98 (0.94-1.03) |  |  |
| C-reactive protein (log scale) | 1.22 (0.99-1.51) |  |  |
| D-dimer > 1mg/dL FEU | 1.23 (0.92-1.64) |  | 2.79 (1.53-5.09)* |
| Ferritin | 1.1 (0.95-1.27) |  |  |
| Troponin detected | 1.89 (1.35-2.64)* |  | 2.49 (1.5-4.14)* |

†Baseline physiologic features and lab findings include the first recorded value assessed within the first 48 hours of admission. Missing values were imputed using multiple imputation prior to model creation. ‡The outcome is peak World Health Organization COVID-19 ordinal disease severity scale ≥5 corresponding to a composite of death or requirement of supplemental oxygen exceeding nasal cannula reported for all patients and patients < 60 years. \*p < 0.05.

A composite severe illness or death outcome was associated with increasing BMI (aHR 1.19 per 5 unit increase; 95%CI 1.11-1.28), self-reported respiratory symptoms (aHR 2.11; 95%CI 1.13-3.94), respiratory rate >18 (aHR 1.12 per increase of 1 over 18; 95%CI1.1-1.15), fever >38.4° Celsius (aHR 1.77; 95%CI 1.36-2.32), elevated troponin (aHR 1.89; 95%CI 1.35-2.64) and albumin (aHR 1.30 per 0.5 g/dL decrease; 95%CI 1.15-1.47) (**Table 2**). A sensitivity landmark analysis excluding those who achieved an outcome within 6 hours of admission is shown in **Table S4**. Most associations were similar to the primary analysis but magnitudes of associations with respiratory and constitutional symptoms weakened. In a sub-group analysis of patients < 60 years of age, we identified male sex (aHR 1.7;95%CI 1.11-2.58), BMI (aHR 1.25 per 5-unit increase; 95%CI 1.14-1.37), CCI (aHR 1.27; 95%CI 1.1-1.46) and respiratory rate (aHR 1.16 per increase of 1 over 18; 95%CI 1.13-1.2) as significantly associated with severe illness or death (**Table 2, Table S5**).

## DISCUSSION

Our study provides valuable insight into the disease trajectories of hospitalized COVID-19 patients in the US and the risk factors associated with severe outcomes. Patients who developed severe illness and survived had a median length of stay of 15 days with 25% having a 20 day stay or longer. We observed an overall mortality of 14%, with nearly half of all deaths occurring among nursing home residents, many of whom had DNI/DNR orders on admission. Although >60% of patients were non-white, we did not observe statistically significant race/ethnicity associations. Obesity and overall comorbidities were significantly associated with severe illness or death, particularly in persons younger than 60 years. Lastly, we found a few simple-to-measure markers such as respiratory rate, D-dimer, and troponin to be strongly associated with death. Knowledge about disease trajectory and outcome is critical as providers, health systems and public health agencies plan for potentially scarce resources such as ventilators and therapeutics.^20,21^ It is also important to understand disease trajectory to determine appropriate levels of care, and when discussing goals of care with patients and families.

Increasing age was strongly associated with death; each decade increase had a 54% increased hazards of mortality. Similar age associations are now well described.^22,23^ Nursing home residents had a 2.1 fold increased hazards of death independent of age or comorbidity score, illustrating the vulnerability of this population to SARS-CoV-2.^24^ There are approximately 1.4 million nursing home residents in the US.^25^ It is estimated that one-third of US COVID-19 deaths are among this population. We found that 49% of deaths occurred in nursing home residents, similar to the 48% of deaths in this population that have been reported in Maryland.^26^ Many of our older patients, including nursing home residents, had advanced medical directives and were DNR/DNI. This clearly impacted the level of intervention, measurement of severity (e.g. lack of mechanical ventilation) and time to death. The implementation of advanced directives varies substantially globally as does the age of the population.^27^ Some of the global differences in SARS CoV-2 mortality are likely due to such differences.

Persons under 60 years of age comprised only 8% of deaths (9 cases) but accounted for the majority of severe illness outcomes among those who were discharged (95 cases; 82%). More than half of those with severe illness were obese, >60% were non-white, and 57% were male. We found that increasing BMI, CCI and male sex but not race/ethnicity were strongly associated with severe disease in those <60 years. The age-adjusted prevalence of obesity in the adult US population is 42.4%. Obesity prevalence is higher among African Americans and Hispanics and linked to socioeconomic status, other comorbidities, and poor health outcomes.^28^ It is not surprising that that there is strong association between high BMI and severe COVID-19 in the US, particularly in younger age groups who are more likely to be obese.^29^ The association between obesity and poor COVID-19 outcomes has also been reported internationally^30,31^ but the causal link remains unknown. Mechanics of breathing may be impaired in obesity. Inflammation caused by excessive fat cells might worsen the hyperinflammatory response seen in COVID-19.^32^ Future studies of COVID-19 interventions should include this vulnerable population.

We found that a few simple-to-measure, baseline laboratory markers, namely absolute lymphocyte count (ALC), albumin, D-dimer and troponin, were associated with progression to severe disease or death. Lymphopenia is highly prevalent in COVID-19, but its impact on mortality has been inconsistent across cohorts.^6^ In our cohort lymphopenia was associated with increased illness severity or death, but not death alone. Low albumin was also associated with severe disease or death, a finding that has been previously observed.^33^ Both elevated D-dimer and troponin were associated with a nearly 3-fold increased risk of death. An elevated D-dimer is associated with increased risk of death in COVID-19 patients independent of documented thromboembolic disease, but could also indicate an increased risk of thrombosis.^34^ An elevated troponin was also an important factor in models of severe disease or death. Whether SARS-CoV-2 leads to direct or indirect cardiac toxicity, it is clear that there is a link between cardiac injury and severe outcomes in COVID-19.^35^

The presence of respiratory symptoms was associated with severe disease while the presence of constitutional symptoms seemed to be protective. This suggests that there are distinct phenotypes in COVID-19 that confer differential risk. Magnitudes of associations with respiratory and constitutional symptoms weakened in our landmark analysis potentially highlighting these symptoms as strong indicators of disease severity upon admission.

Lastly, we found that an elevated respiratory rate was associated with severe outcomes. This likely reflects that severity of illness in COVID-19 is tightly linked to pulmonary complications including acute respiratory distress syndrome (ARDS) and thromboembolism. Respiratory rate is included in mortality prediction scores for hospitalized patients^36,37,^ and has been associated with mortality in COVID-19.^38^ The fact that such an easily measured parameter associates more strongly with severe outcome than several inflammatory markers (e.g. CRP, ferritin) suggests that inexpensive and immediately available metrics can provide valuable information about disease trajectory.

There are some limitations to our study. Ten percent of the patients in our cohort did not yet have an observed outcome and such incompleteness could lead to bias. However, since we adopted time-to-event approaches which handled censored survival data, our analyses remain unbiased and not affected by incompleteness. Our data are derived from a single health system and may not be representative of COVID-19 populations across the US. Care practices may have differed between our 5 hospitals. We may have under ascertained the number of COVID-19 positive cases in our health system due to testing challenges.^39^ We may not have captured all comorbidities since some patients may not have had robust documentation in the electronic health record. Post-discharge outcomes are not currently captured if they occur outside of the health system. Lastly, we had to impute a considerable percentage of missing values in several laboratory tests as there is no clear standard of care for laboratory testing in COVID-19.

In conclusion, we identified several important demographic and simple to assess factors associated with severe COVID-19 outcomes including age, nursing home status, BMI, D-dimer, troponin, ALC and respiratory rate. We also identified specific subgroups with a higher risk of disease progression including the elderly, nursing home residents, and younger patients with obesity.

## Data Availability

The data for this study is maintained in the Johns Hopkins Precision Medicine Analytics Platform (PMAP).

## ACKNOWLEDGEMENTS

The data utilized for this publication were part of the JH-CROWN: The COVID PMAP Registry, which is based on the contribution of many patients and clinicians. JH-CROWN received funding from Hopkins inHealth, the Johns Hopkins Precision Medicine Program.

## Supplemental Appendix: Patient trajectories and risk factors for severe COVID outcomes among persons hospitalized for COVID-19 in the Maryland/DC region

### Natural Language Processing

In order to identify symptoms at presentation, we first created a meta-lexicon of four symptom categories (organized into 11 sub-categories) based on the guidelines provided by the CDC, WHO, and clinical findings. For each symptom category, we generated a set of synonym terms using the Unified Medical Language System (UMLS) Metathesaurus,^1^ and we iteratively worked with domain experts to revise the symptom categories and synonyms. Table S4 includes the list of symptom categories and the search terms.

We then selected relevant clinical note types for each patient, including H&P, Critical Care Notes, Progress Notes, and ED Notes, focusing specifically on the notes created within 48 hours before and after admission. Next, we pre-processed the note text and extracted only the relevant narrative parts, particularly the chief complaint and history of the present illness sections.

We then used a COVID-19-customized version of MedTagger,^2^ together with our in-house Python tools to (a) identify phrases and synonyms of particular symptoms within the text narratives, (b) determine if these symptom mentions are negated, possible, or positive in their context, (c) classify symptoms into the predefined 11 categories, and (d) map them to their corresponding UMLS Concept Unique Identifiers (CUIs). These NLP pipelines use a combination of machine learning models, including Conditional random fields (CRFs),^3^ and contextual rule-based methods, including regular expressions. Finally, we selected only the positive symptom mentions in the notes and aggregated all presenting symptoms for each patient.

To evaluate the performance of our NLP methods, two abstractors manually reviewed over 100 notes from 20 randomly selected patients. For each patient, each symptom was labeled as present or not-present (same label set as the NLP output), resulting in 220 manually labeled symptoms with the inter-rater agreement of 97%. The 3% disagreements were individually adjudicated between the two abstractors. Comparing the created gold standard to the labels generated by the NLP methods, we found that we could achieve the following results:

| <u>Metric</u> | <u>Value (95% CI)</u> |
| --- | --- |
| Sensitivity: | 0.956 (0.913 – 0.998) |
| Specificity: | 0.931 (0.887 – 0.974) |
| Negative Predictive Value: | 0.968 (0.937 – 0.999) |
| False Positive Rate: | 0.069 (0.026 – 0.113) |
| False Negative Rate: | 0.044 (0.002 – 0.087) |

**Table S1.**
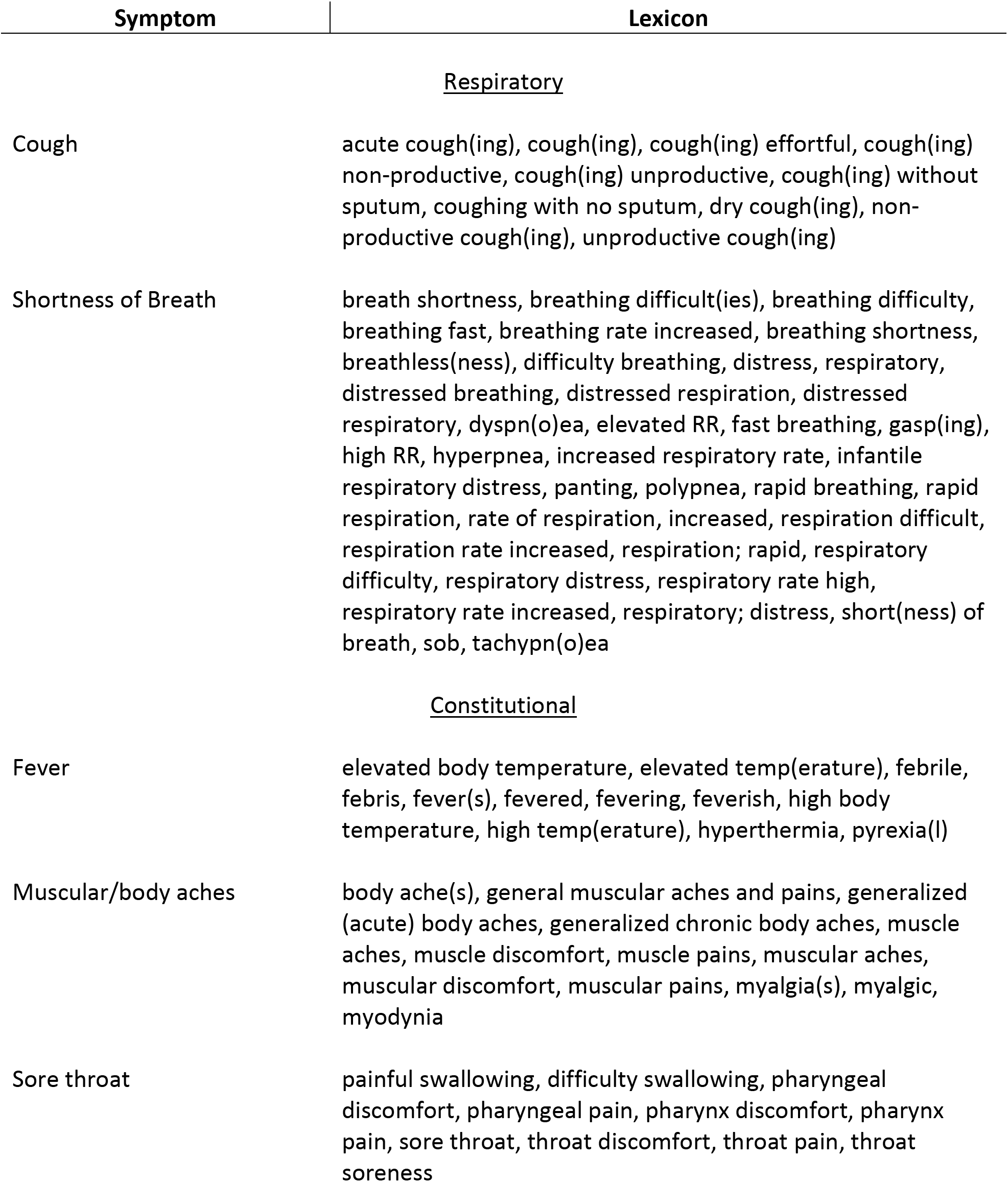
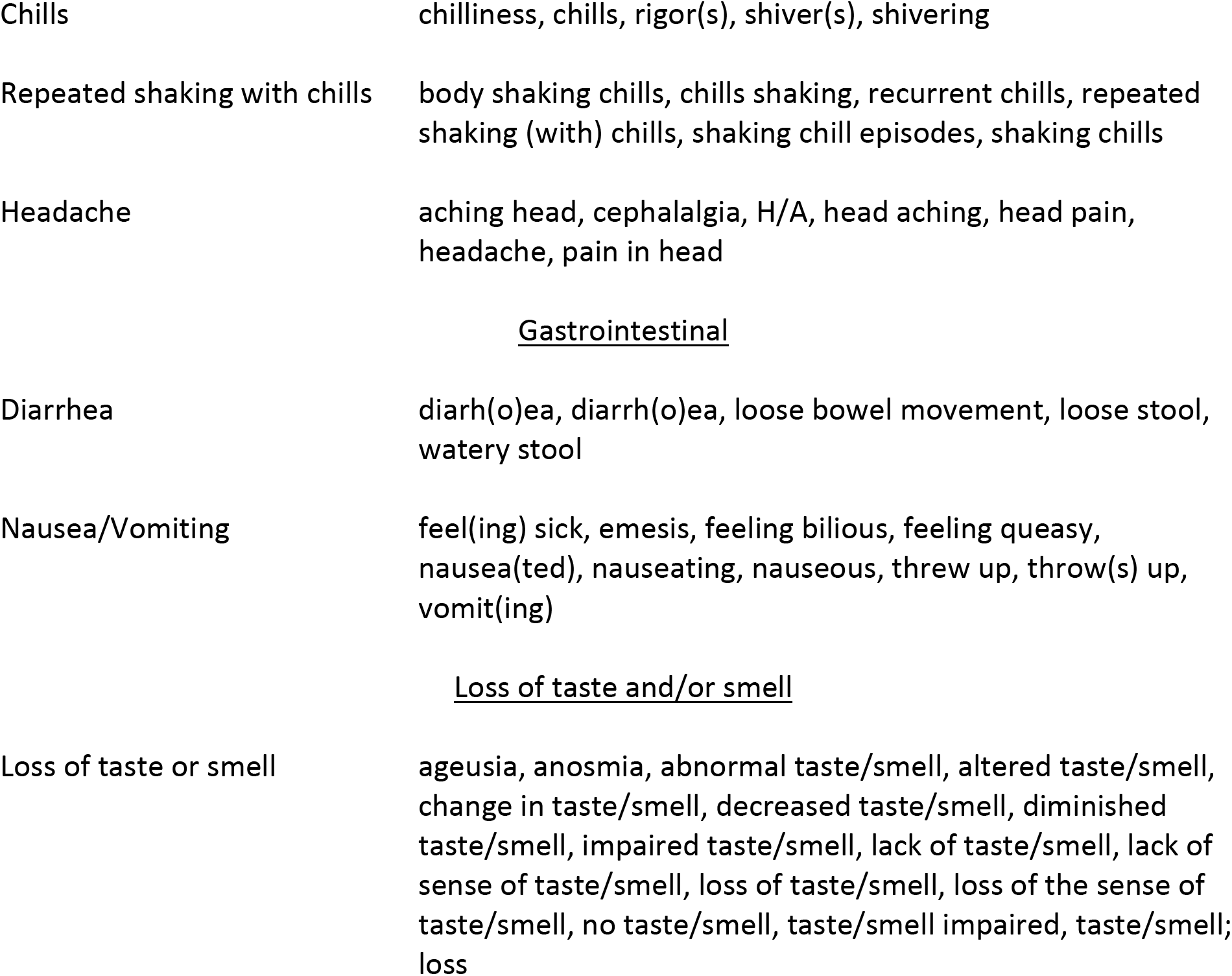
Meta-lexicon of symptom categories and synonym terms for NLP text mining

**Table S2.** Medications administered in the hospital to patients with observed outcomes

| Medication | Outcome Observed (n=748) |  |  |  |
| --- | --- | --- | --- | --- |
|  | Overall<br>(N = 832) | Mild/Moderate,<br>(N = 518) | Severe,<br>N = 116 | Death,<br>N = 113 |
| ACE inhibitors | 80 (10%) | 46 (9%) | 27 (14%) | 7 (6%) |
| Antibiotics | 637 (76%) | 355 (67%) | 176 (94%) | 106 (93%) |
| Antifungals | 63 (8%) | 11 (2%) | 33 (18%) | 19 (17%) |
| ARBs | 71 (9%) | 50 (9%) | 17 (9%) | 4 (4%) |
| Baricitinib | 0 (0%) | 0 (0%) | 0 (0%) | 0 (0%) |
| Corticosteroids | 121 (15%) | 41 (8%) | 51 (27%) | 29 (25%) |
| DAS-181 or placebo<sup>a</sup> | 6 (1%) | 3 (1%) | 2 (1%) | 1 (1%) |
| Heparin or enoxaparin<sup>b</sup> | 571 (69%) | 366 (69%) | 146 (78%) | 59 (52%) |
| Hydroxychloroquine | 384 (46%) | 217 (41%) | 103 (55%) | 64 (56%) |
| Ibrutinib | 1 (0%) | 1 (0%) | 0 (0%) | 0 (0%) |
| Oseltamivir | 2 (0%) | 1 (0%) | 1 (1%) | 0 (0%) |
| Remdesivir<sup>a</sup> | 8 (1%) | 4 (1%) | 4 (2%) | 0 (0%) |
| Ritonavir | 1 (0%) | 0 (0%) | 1 (1%) | 0 (0%) |
| Rituximab | 1 (0%) | 0 (0%) | 1 (1%) | 0 (0%) |
| Statins | 271 (33%) | 160 (30%) | 68 (36%) | 43 (38%) |
| Tocilizumab | 39 (5%) | 4 (1%) | 29 (16%) | 6 (5%) |

<sup>a</sup> Patients were enrolled in a randomized trial. <sup>b</sup>Heparin and enoxaparin does not distinguish between prophylactic or therapeutic dosing.

**Table S3:** Association of clinical characteristics and mortality with SaO2/FiO2 included

| Clinical characteristic<sup>†</sup> | Death, hazard ratio (95% CI) |
| --- | --- |
| Age (increments of 10 years) | 1.66 (1.36-2.02)* |
| Admission from nursing home | 2.16 (1.41-3.32)* |
| Male sex | 1.12 (0.74-1.72) |
| Body mass index (increments of 5 kg/m<sup>2</sup>) | 1.08 (0.95-1.23) |
| Non-white race/ethnicity | 0.84 (0.54-1.31) |
| Charlson comorbidity index | 1.14 (1.02-1.27)* |
| Respiratory rate < 17 breaths per minute | 0.7 (0.49-1.01) |
| Respiratory rate 17-25 breaths per minute | 1.13 (1.02-1.25)* |
| Respiratory rate > 25 breaths per minute | 1.06 (1-1.13) |
| SaO<sub>2</sub>:FiO<sub>2</sub> < 375 (decreasing increments of 75) | 1.35 (1.16-1.56)* |
| SaO<sub>2</sub>:FiO<sub>2</sub> ≥ 375 (decreasing increments of 75) | 0.94 (0.65-1.39) |
| D-dimer > 1mg/dL FEU | 2.48 (1.36-4.54)* |
| Troponin detected | 2.24 (1.35-3.73)* |

<sup>†</sup>Baseline physiologic features and lab findings include the first recorded value assessed within the first 48 hours of admission. Missing values were imputed using multiple imputation prior to model creation. \*p < 0.05

**Table S4:** Factors associated with severe disease and/or death excluding patients who achieved those outcomes in the first six hours of admission

| Clinical characteristic<sup>†</sup> | Severe outcome or death, hazard ratio (95% CI) <sup>‡</sup> |
| --- | --- |
| Age (increments of 10 years) | 1 (0.87-1.16) |
| Admission from nursing home | 1.14 (0.71-1.83) |
| Age-nursing home interaction term | 0.92 (0.71-1.17) |
| Male sex | 0.92 (0.66-1.28) |
| Body mass index (increments of 5 kg/m2) | 1.23 (1.12-1.36)* |
| Non-white race/ethnicity | 1 (0.71-1.42) |
| Charlson comorbidity index | 1.03 (0.92-1.15) |
| Respiratory symptoms | 1.4 (0.71-2.75) |
| Gastrointestinal symptoms | 1.03 (0.75-1.43) |
| Constitutional symptoms | 1.26 (0.62-2.55) |
| Loss of taste and/or smell | 0.94 (0.58-1.53) |
| Respiratory rate < 18 breaths per minute | 0.81 (0.63-1.03) |
| Respiratory rate > 18 breaths per minute | 1.12 (1.09-1.16)* |
| Pulse (increments of 10) | 1.05 (0.95-1.16) |
| Fever (Temperature >38.4o Celcius) | 1.63 (1.17-2.27)* |
| Absolute lymphocyte count (log scale) | 0.71 (0.53-0.96)* |
| Alanine aminotransferase (log scale) | 1 (0.8-1.24) |
| C-reactive protein (log scale) | 1.19 (0.92-1.54) |
| D-dimer > 1mg/dL FEU | 1.01 (0.69-1.47) |
| Ferritin | 1.18 (0.98-1.41) |
| Troponin detected | 1.7 (1.12-2.57)* |

<sup>†</sup>Baseline physiologic features and lab findings include the first recorded value assessed within the first 48 hours of admission. Missing values were imputed using multiple imputation prior to model creation.
<sup>‡</sup>The outcome is peak World Health Organization COVID-19 ordinal disease severity scale ≥5 corresponding to a composite of death or requirement of supplemental oxygen exceeding nasal cannula.\*p < 0.05

**Table S5:** Association of clinical characteristics and severe outcomes, age < 60, demographics not forced into model

| Clinical characteristic<sup>†</sup> | Severe outcome or death, hazard ratio (95% CI)<sup>‡</sup> |
| --- | --- |
| Body mass index (increments of 5 kg/m<sup>2</sup>) | 1.18 (1.08-1.3)* |
| Charlson comorbidity index | 1.12 (0.95-1.31) |
| Respiratory rate < 18 breaths per minute | 0.78 (0.62-0.99)* |
| Respiratory rate > 18 breaths per minute | 1.11 (1.07-1.15)* |
| Albumin (every increase of 0.5 g/L) | 0.86 (0.73-1.01) |
| Glomerular filtration rate (increments of 10 ml/min) | 0.99 (0.92-1.05) |
| C-reactive protein (log scale) | 1.72 (1.27-2.33)* |
| Troponin detected | 1.93 (1.07-3.45)* |

<sup>†</sup>Baseline physiologic features and lab findings include the first recorded value assessed within the first 48 hours of admission. Missing values were imputed using multiple imputation prior to model creation. <sup>‡</sup>The outcome is peak World Health Organization COVID-19 ordinal disease severity scale $\geq 5$ corresponding to a composite of death or requirement of supplemental oxygen exceeding nasal cannula. \*p < 0.05

